# Digital phenotyping using wearable-determined physical behaviors and machine learning to detect depression and anxiety in a general population

**DOI:** 10.1101/2025.09.01.25334782

**Authors:** Alireza Sameh, Laura Nauha, Marjo Seppänen, Maisa Niemelä, Mourad Oussalah, Raija Korpelainen, Vahid Farrahi

## Abstract

**Objective:** Depression and anxiety are widespread mental health disorders, yet their diagnosis remains challenging. Digital phenotyping with wearable devices provides a promising approach for detecting depression and anxiety in the general population. This study aims to explore the extent to which wearable accelerometer-determined physical behavior metrics can be used as digital phenotypes for identifying individuals with and without depression and anxiety symptoms using machine learning (ML) algorithms.

**Methods:** At age 46 years old, participants (N = 2,810) from the Northern Finland Birth Cohort 1966 carried wrist- and waist-worn accelerometers for 14 consecutive days. Physical activity and sedentary behaviors were measured using data from the waist-worn device, while sleep behavior was identified based on data from the wrist-worn accelerometer. A total of 54 physical behavior metrics were extracted for each participant. Severity of the depression and anxiety symptoms were assessed using three validated instruments: the Beck Depression Inventory-II, Generalized Anxiety Disorder-7, and the Hopkins Symptom Checklist-25. Five ML algorithms were applied to identify individuals with and without depression and anxiety symptoms. Model interpretability was enhanced using SHapley Additive exPlanations (SHAP) to assess the contribution of individual features.

**Results:** Among ML models, random forest achieved the best performance with accuracy (66%–72%) and AUC (66%–70%) for all three instruments. Physical behavior metrics extracted from accelerometers emerged as potential predictors of depression and anxiety. In SHAP analysis wake up time, time in bed, bed time, physical activity intensity proportions and prolonged sedentary bouts emerged as most important features.

**Conclusions:** Wearable-derived metrics of physical behaviors combined with ML models can be utilized with reasonably good accuracy to differentiate between participants with and without depression and anxiety symptoms. Our findings support the utility of wearable-derived physical behavior digital phenotypes for differentiating between participants with and without depression and anxiety symptoms in a general population.

## 1. INTRODUCTION

Depression and anxiety have become more prevalent, placing immense pressure on healthcare systems^1,2^. Depression is characterized by persistent low mood, loss of interest in activities, exhaustion, cognitive difficulties, feelings of guilt or worthlessness, and social withdrawal, while anxiety disorders often feature excessive worry, restlessness, and heightened alertness^3^. Many individuals experience a mix of both, sharing symptoms such as sleep disturbances, excessive rumination, negative self-image, trouble focusing, and, in severe cases, suicidal thoughts^4,5^. These conditions, influenced by factors like age, gender, socioeconomic status, environment, and lifestyle, can range from mild to severe and significantly affect daily functioning, interpersonal relationships, and overall quality of life^6^.

Traditional and clinical methods for assessing depression and anxiety such as structured interviews and self-reported methods remain essential. However, these approaches have notable limitations^7^. Assessment of depression and anxiety symptoms using interviews and self-reported methods rely on retrospective recall, which can introduce bias and fail to reflect the dynamic nature of mental health symptoms including depression and anxiety as they unfold in daily life^8^. Additionally, these methods have generally limited capacity for early detection of depression and anxiety, as they typically require clinical visits and access to expert opinions. Timely identification is important for a number of reasons, including enabling access to appropriate support, preventing symptom progression, and improving overall quality of life^7^. Recent research highlights a need for collecting data in real life settings to obtain more accurate and ecologically valid insights into the patterns and signs of depression and anxiety symptoms, enabling earlier and more precise detection^9,10^.

With the widespread adoption and availability of personal digital devices, data generated by these devices can be utilized to identify phenotypic features and indicators, referred to as “digital phenotypes,” that are applicable for assessing and identifying various mental disorders^11^. Digital phenotyping offers a promising approach to addressing challenges in mental health by leveraging continuous, moment-by-moment digital data captured through personal devices in the context of individuals’ daily lives. Digital phenotyping holds particular potential for enabling early, rapid, and accurate detection and screening of mental disorders^11^. To date, digital phenotyping of depression and anxiety have been most frequently performed by utilizing different types of data and sensing signals collected from smartphones, often in combination with machine learning (ML) methods^11,12^. These data and signals often include mobility patterns (via GPS), movement patterns (via accelerometer), social networks and social dynamics (via call and text logs and Bluetooth), and voice samples (via microphone)^13^. For instance, changes in mobility patterns tracked via GPS such as reduced daily travel distance or time spent at home have been related to depressive symptoms^14,15^. Similarly, phone usage metrics, including decreased communication frequency (e.g., fewer calls or texts) and disrupted sleep inferred from screen activity, have shown correlations with both depression and anxiety^14,16^.

Among the multitude of digital phenotypes derived from different smartphone signals and data for identifying individuals at high risk of depression and anxiety, preliminary evidence suggests that depression and anxiety may particularly manifest in digital phenotypes of movement patterns derived from smartphone-based accelerometer sensors^11,17^. A preliminary, hypothesis-generating study previously showed that social anxiety disorder symptom severity can be predicted by smartphone-collected data, with the most important features being movement patterns identified from accelerometer sensors, rather than social features such as calls or texts^18^. This finding is in line with the existing literature suggesting that depression and anxiety symptoms are linked to physical behaviors such as lower level of physical activity, higher level of sedentary behaviors, and sleep problems^19–21^.

The extent of digital phenotyping has recently expanded towards signals collected from wearable devices such as wearable activity and fitness trackers, and heart rate monitors. Compared to smartphones, data collected from wearable devices may have several advantages for the applications of digital phenotyping^22^. Wearable devices are typically worn at a fixed body location, which allows for high-quality physiological and behavioral data, such as temperature, heart rate, and physical behaviors^23,24^. Additionally, the continuous wearing of these devices allows for more consistent and uninterrupted measurement of signals over time^25,26^.

Modern research-grade and consumer-grade wearable devices can monitor a wide range of physiological parameters, including heart rate, skin temperature, motion, and electrodermal activity^10,23,27^. Despite incorporating multiple sensing modalities, accelerometry remains a foundational technology present in most wearable devices. Accelerometer data from wearable devices, whether multisensory or focused only on activity, are widely used in research primarily for long-term, accurate, and moment-by-moment monitoring of physical behaviors including physical activities, sedentary behaviors, and sleep and other aspects of these behaviors such as rest-activity rhythm metrics, accumulation patterns, and temporal distribution^28,29^.

With the advancement of ML, several studies have examined the use of continuous accelerometer data from wearable devices to identify the risk of chronic diseases and health conditions^23,30,31^. This includes efforts to develop scalable and rapid methods for screening individuals at high risk of various mental health outcomes^23^. ML algorithms trained on statistical features derived from wearable accelerometry signals have demonstrated their ability to identify individuals at risk for neurodegenerative diseases such as Alzheimer’s and Parkinson’s diesease^10,32^. Similarly, ML models utilizing metrics of physical activity, sedentary behavior, and sleep have been successfully used to predict outcomes such as mortality risk^33^, type 2 diabetes^34^, and obesity^35^. Recent analysis of wearable accelerometry data from a nationally representative sample of the U.S. general population identified features like quartile-distributed activity levels, mean daily activity across weekdays and weekends, and full-week movement trends as key predictors of major depressive disorder^36^. Similarly, wearable accelerometer-determined indicators of sleep latency, sleep efficiency, and movement intensities have demonstrated predictive capability in identifying early signs of symptom deterioration in individuals with anxiety disorders^37^.

Given the early evidence suggesting that the presence of depression and anxiety may manifest in physical behaviors, further research is needed to examine whether wearable accelerometer-determined physical behavior metrics can be used as digital phenotypes for identifying individuals at high risk of depression and anxiety. The aim of this population-based study was to explore the extent to which wearable accelerometer-determined physical behavior metrics can be used as digital phenotypes for identifying individuals with and without depression and anxiety symptoms using ML algorithms in a general population.

## 2. METHODS

Our study advocates a structured ML pipeline to predict the presence or absence of depression and anxiety symptoms in a general population using physical activity, sedentary behaviors, sleep related features determined from two wearable accelerometers in wrist and waist. The methodology consists of two main stages. First, wearable data were processed to determine physical activity, sedentary behaviors, and sleep period on the basis of previously developed and validated techniques^38–40^. Physical activity and sedentary behaviors were identified from a waist worn accelerometer device and sleep period from a wrist-worn accelerometer-based activity monitor. From the identified physical activity, sedentary behavior, and sleep periods, a total number of 54 features were computed for each participant. The features were computed for weekdays, weekends, and the full week. Depression and anxiety symptoms were assessed by three validated and commonly used screening tools. The extracted features were then used as input features to ML modeling to detect depression and anxiety. The model’s performance is assessed using evaluation metrics, and SHAP analysis is applied to interpret the contribution of each feature. Figure 1 illustrates the overview of the study.

**Figure 1:**
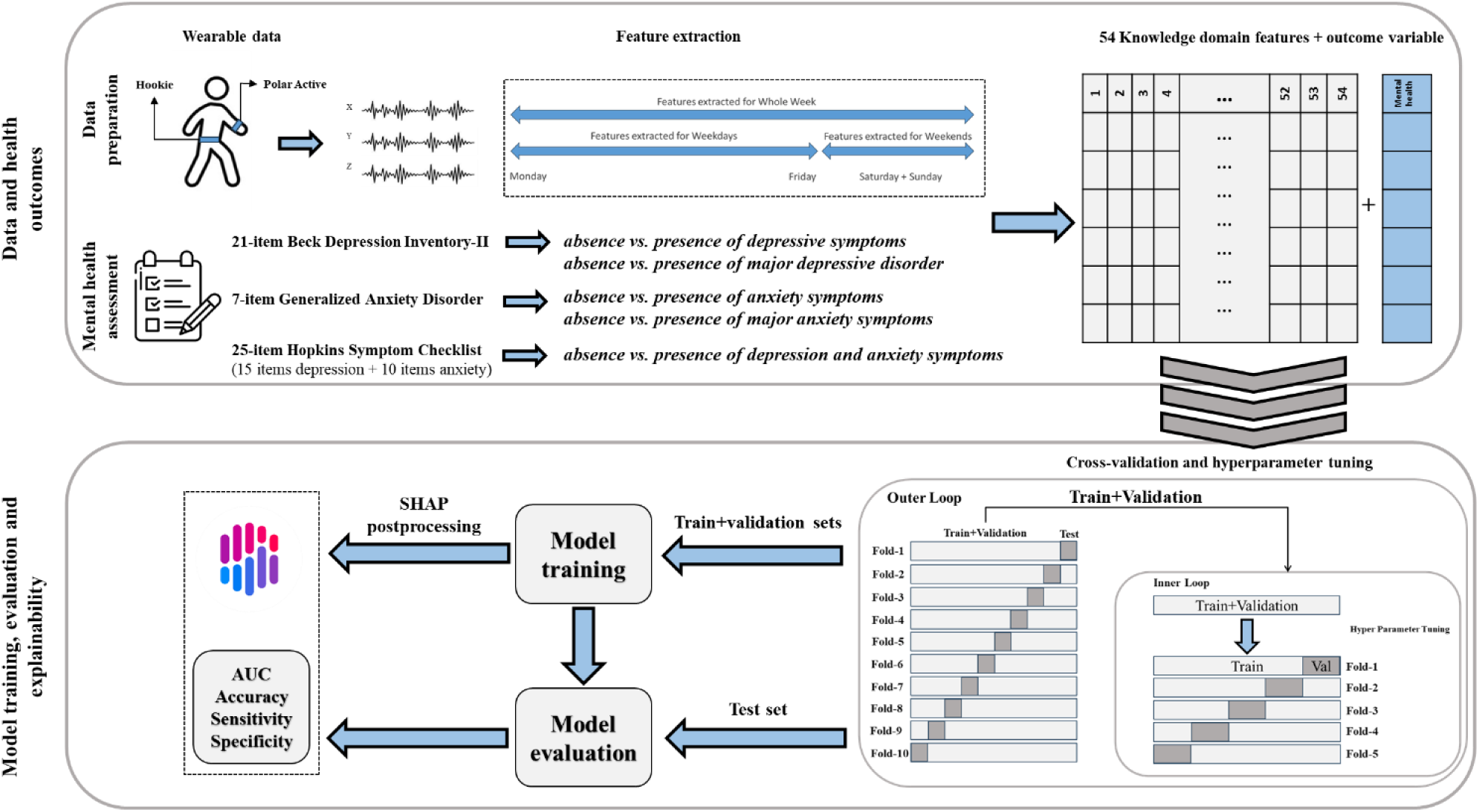
The flow of study consists of two main steps. First, wearable signals and questionnaires are converted to the structured data set to apply machine learning models. Second, machine learning methods were performed and feature explainability anal

### 2.1. Study population

Data for this study was from the population-based Northern Finland Birth Cohort 1966 (NFBC1966) study^41^. NFBC1966 (N = 12,058) is a life-course cohort study involving participants born in 1966 in Northern Finland. Since birth, the cohort members have then been regularly monitored prospectively with a broad set of clinical measurements, interviews, and postal questionnaires. This study analyzed the data from the most recent time point when individuals were 46 years old, with a target population of 10,321. The data collected in the 46-year follow-up included monitoring of daily activities and sleep using two wearable devices (i.e., a waist-worn accelerometer and a wrist-worn accelerometer-based activity monitor), and mental health assessment with three questionnaires. Eligible participants for this study were those cohort members who wore wearable accelerometers, provided enough valid accelerometry data, and completed the questionnaires for mental health assessment. NFBC1966 was approved by the Ethical Committee of the Northern Ostrobothnia Hospital District (94/2011), and written informed consent was obtained from all the participants. More details on the follow-up, including loss of follow-up and missingness, has been described earlier by previous research^42^.

### 2.2. Monitoring of physical activity, sedentary and sleep

Participants were asked to wear simultaneously two accelerometers, one on their waist (Hookie AM20) and one on wrist (Polar Active) for 14 consecutive days. They were instructed to wear the waist-worn accelerometer during all waking hours except water-based activities, and the wrist-worn accelerometer continuously for 24 hours, including sleep time. The Hookie was worn on the waist with an elastic belt and the Polar Active was worn on the wrist of nondominant hand. The Hookie accelerometer is a research-grade device configured to measure triaxial raw acceleration signals at 100 Hz^43^. The Polar Active is a consumer-grade device that provides metabolic equivalents (METs) every 30 seconds, derived from uniaxial acceleration signals combined with background information such as body height, body weight, age, and sex. These estimates are generated using a proprietary built-in algorithm developed by the manufacturer.

Both of these accelerometers have been previously validated for measurement of physical behaviors. Polar Active has demonstrated strong accuracy in detecting energy expenditure during leisure activities (correlation coefficient of 0.88) and exercise protocols, including lower and upper-body strength training and cycling (correlation coefficient of 0.79)^44^. Similarly, the Hookie AM20 has been validated against indirect calorimetry and reference devices, showing high accuracy in estimating physical activity intensity and energy expenditure across both controlled treadmill tasks and free-living conditions (correlation coefficients ranging from 0.86 to 0.89)^45–47^.

### 2.3. Mental health assessment

Depression and anxiety symptoms were assessed using three standardized instruments, including Beck Depression Inventory–II (BDI-II), Generalized Anxiety Disorder 7-item Scale (GAD-7), and the Hopkins Symptom Checklist–25 (HSCL-25). These rating scales have all been widely used as a questionnaire for assessment of depression and anxiety symptoms. BDI-II consists of 21 items; each rated on a scale from 0 to Total scores range from 0 to 63, with higher scores indicating more severe levels of depression^48,49^. The GAD-7 includes 7 items indicating anxiety symptoms, each rated from 0 to 3, resulting in a total score between 0 and 21^50^. The HSCL-25 contains 25 items rated on a 4-point scale, from 1 (“Not at all”) to 4 (“Extremely”). It is divided into subscales for anxiety (10 items) and depression (15 items), and the average score is calculated for assessment^51^.

### 2.4. Wearable-measured physical behaviors (physical activity intensities, sedentary behavior, sleep)

For the purposes of this study, physical activity and sedentary behavior features were extracted from Hookie, and sleep features from Polar Active. We used the waist worn accelerometer data to identify physical activity and sedentary behavior features, because the waist is closer to center of body. Previous studies have shown that waist-worn accelerometry provides a relatively more accurate estimation of physical activity and sedentary behaviors^52^. However, since Hookie device was worn only during waking hours, sleep period could not be detected from this device. We identified sleep behavior from wrist-worn Polar Active, which was worn also during sleeping^38^. To be eligible for inclusion in the study, participants were required to provide seven consecutive valid days (one full week) of accelerometer data from both the Hookie and Polar Active devices. In agreement with prior studies^39,53^, a valid measurement day was defined as having at least 10 hours of wear time for Hookie device. A valid measurement day for Polar Active was defined as having valid measurement with no less than 2.5 hours of non-wear time.

#### 2.4.1. Physical activity and sedentary behaviors detection from waist worn accelerometer

Physical activity and sedentary behaviors were determined from raw waist-worn accelerometry (Hookie device). Raw signals were segmented into 6-second windows and mean amplitude deviation (MAD) values were computed for each window. MAD is a widely used method for analyzing raw accelerometer data and measurement of physical activity intensities and sedentary behaviors^54^. From the 6-s MAD values, monitor non-wear time was detected and removed using a previously validated method^55^. A non-wear time period was defined as ≥ 90 consecutive minutes of no detected movement, allowing for short movement intervals of up to 30 seconds, if no other movements were detected in the 30 min either side of the current 30-min interval^56^. The remaining time-epochs were then marked as sedentary behavior (<1.5 metabolic equivalents [MET]), light physical activity (LPA)(1.5 – 3.0 MET), or moderate-to-vigorous physical activity (MVPA)(≥ 3MET) using a validated set of thresholds for MAD values^46^.

#### 2.4.2. Sleep behavior detection from wrist worn accelerometer

Participant’s time in bed was detected using a previously validated algorithm, which identifies bedtime and wake-up time based on analyzing MET-values. The algorithm is designed to identify bedtime and wake up time by analyzing active and inactive periods from Polar Active output MET-values^57,58^. Briefly, the algorithm separates MET values over consecutive 24-hour windows, from 18:00:00 to 17:59:30 the following day. Within each 24-hour period, all sustained movement bouts (defined by continuous MET values ≥1 lasting more than 45 minutes) are identified. The algorithm then examines the intervals between these sustained bouts to detect potential sleep periods. This algorithm was validated by comparing device-estimated sleep intervals against self-reported sleep diaries. In a study involving 108 adults, the algorithm demonstrated strong agreement with self-reported bedtimes and wake-up times, achieving intraclass correlation coefficients (ICCs) greater than 0.81 for both metrics. Mean differences between device-estimated and self-reported bedtimes were less than 6 minutes, and for wake-up times, less than 19 minutes^38^.

### 2.5. Machine learning modeling for depression and anxiety detection

#### 2.5.1 Feature extraction

This study extracted a comprehensive set of physical activity, sedentary behaviors, and sleep features grounded in prior research indicating their significance in association with different health outcomes. These features were computed separately for weekdays, weekend days, and ^36,59^whole week to capture differences between weekday and weekend days, and to reflect overall weekly patterns^36^. Table 1 presents the full list of extracted features and their description. The physical activity and sedentary behavior features included measures of intensity distribution and accumulation patterns. Intensity distribution features encompassed the proportions of wearing time spent in sedentary behavior (<1.5 METs), LPA, and MVPA, each normalized to total accelerometer wear time. We also computed active time proportion as the combined proportion in LPA and MVPA. Total energy expenditure was expressed as average MET-hours per day. Sedentary bout characteristics, quantifying frequency and cumulative duration across predefined bout durations (0–5, 5–10, 10–15, 15–30, ≥30 min) were also calculated. Sleep-related features included the mean wake-up time, bedtime, and total time in bed, each averaged across the entire week, as well as separately for weekdays and weekends. Wake-up time and bedtime were computed as the mean time (in minutes from midnight) across days in each time segment. The sleep period, labeled as time in bed was calculated as the duration between bedtime and wake-up time.

**Table 1:** The name and description of 54 features extracted based on both (waist+wrist) accelerometers data.

| Feature names | Description |
| --- | --- |
| **Polar Active** |  |
| Whole week wake up time | Mean of wake-up time across whole week |
| Weekdays wake up time | Mean of wake-up time across weekdays |
| Weekend wake up time | Mean of wake-up time across weekend |
| Whole week bedtime | Mean of bedtime across whole week |
| Weekdays bedtime | Mean of bedtime across weekdays |
| Weekend bedtime | Mean of bedtime across weekend |
| Whole week time in bed | Mean of time in bed across whole week |
| Weekdays time in bed | Mean of time in bed across weekdays |
| Weekend time in bed | Mean of time in bed across weekend |
| **Hookie** |  |
| Whole week proportion of sedentary time | Proportion of daily sedentary time from total daily wear time, averaged across whole week |
| Weekdays proportion of Sedentary time | Proportion of daily sedentary time from total daily wear time, averaged across weekdays |
| Weekend proportion of Sedentary time | Proportion of daily sedentary time from total daily wear time, averaged across weekend |
| Whole week proportion of MVPA time | Proportion of daily MVPA time from total daily wear time, averaged across whole week |
| Weekdays proportion of MVPA time | Proportion of daily MVPA time from total daily wear time, averaged across weekdays |
| Weekend proportion of MVPA time | Proportion of daily MVPA time from total daily wear time, averaged across weekend |
| Whole week proportion of LPA time | Proportion of daily LPA time from total daily wear time, averaged across whole week |
| Weekdays proportion of LPA time | Proportion of daily LPA time from total daily wear time, averaged across weekdays |
| Weekend proportion of LPA time | Proportion of daily LPA time from total daily wear time, averaged across weekend |
| Whole week proportion of active time | Proportion of daily LPA+MVPA time from total daily wear time, averaged across whole week |
| Weekdays proportion of active time | Proportion of daily LPA+MVPA time from total daily wear time, averaged across weekdays |
| Weekend proportion of active time | Proportion of daily LPA+MVPA time from total daily wear time, averaged across weekend |
| Whole week MET.Hour | Mean of MET. Hours across whole week |
| weekdays MET.Hour | Mean of MET. Hours across weekdays |
| Weekend MET.Hour | Mean of MET. Hours across weekend |
| Whole week frequency of (0-5, 5-10, 10-15, 15-30, $\geq 30$) min sedentary bout | number of (0-5, 5-10, 10-15, 15-30, $\geq 30$) min sedentary bout averaged across whole week |
| Weekdays frequency of (0-5, 5-10, 10-15, 15-30, $\geq 30$) min sedentary bout | number of (0-5, 5-10, 10-15, 15-30, $\geq 30$) min sedentary bout averaged across weekdays |
| Weekend frequency of (0-5, 5-10, 10-15, 15-30, $\geq 30$) min sedentary bout | number of (0-5, 5-10, 10-15, 15-30, $\geq 30$) min sedentary bout averaged across weekend |
| Whole week duration of (0-5, 5-10, 10-15, 15-30, $\geq 30$) min sedentary bout | Total duration of (0-5, 5-10, 10-15, 15-30, $\geq 30$) min sedentary bout averaged across whole week |
| Weekdays duration of (0-5, 5-10, 10-15, 15-30, $\geq 30$) min sedentary bout | Total duration of (0-5, 5-10, 10-15, 15-30, $\geq 30$) min sedentary bout averaged across weekdays |
| Weekend duration of (0-5, 5-10, 10-15, 15-30, $\geq 30$) min sedentary bout | Total duration of (0-5, 5-10, 10-15, 15-30, $\geq 30$) min sedentary bout averaged across weekend |

*Abbreviations: Metabolic Equivalent of Task; MET, Light Physical activity; LPA, Moderate to Vigorous Physical Activity; MVPA*

#### 2.5.2. Outcome variables

Depression and anxiety symptoms were considered as outcome variables and they were measured using BDI-II, GAD-7, and HSCL-25. Those having a total score of 14 or higher on the BDI-II were considered as having at least mild depressive symptoms^49^, while the BDI-II scores of 20 or higher were considered as having clinically significant depression symptoms^48^. A total score of 5 or higher on the GAD-7 has been commonly used as the threshold for identifying generalized anxiety symptoms, and the total score of 10 and above has been considered as clinically significant generalized anxiety symptoms^50^. These thresholds of GAD-7 were used to classify participants by severity of anxiety symptoms. In this study, the mean score of 1.75 or higher in the HSCL-25 was considered to indicate clinically significant depression and anxiety^51^.

#### 2.5.3. Outlier elimination

One major challenge in using wearable-derived data is the problem of susceptibility to noise, which can be caused by sensor errors, user behavior, and environmental conditions^60,61^. Such noise can introduce outliers that accordingly influence the accuracy and interpretability of ML models^62,63^. Therefore, robust preprocessing methods are important to improve data quality and ensure reliable model performance^63^. After we extracted features, we screened each feature with the Interquartile Range (IQR) rule^64^. For every feature we computed the first quartile (Q1, 25th percentile) and the third quartile (Q3, 75th percentile) and calculated the IQR measure representing the middle 50% of the distribution. Any observation with at least one feature below Q1 − 1.5 × IQR or above Q3 + 1.5 × IQR was labelled an outlier and removed^64^. This non-parametric rule is robust to skew, non-normal data and helps ensure that extreme values do not bias subsequent modelling^64^.

#### 2.5.4. Machine learning models

We employed five commonly used ML algorithms to predict the presence of depression and/or anxiety symptoms. At present, it remains unclear which ML algorithms perform best in predicting health outcomes from wearable sensing data^12,65^. Previous studies have investigated various algorithms in this context, often favoring tree-based methods, likely due to their superior performance and efficiency^12^. Consistent with prior research, we selected Random Forest (RF)^66^, Extreme Gradient Boosting (XGB)^67^, Gradient Boosting Tree (GBT)^68^, and Light Gradient Boosting Machine (LightGBM)^69^ to predict the present of depression and/or anxiety symptoms. In addition to this, we also used Logistic Regression (LR), as it is well-suited for binary classification problems.

We also used a stacked ensemble approach. The stacked ensemble combined RF, XGB, GBT, and LightGBM as base learners. The individual predictions were then integrated using a stacked learning framework to generate the final prediction. This means that the outputs of the base models were used as input features for a second-level model (meta-learner), which could determine how to best combine them to improve overall prediction accuracy.

#### 2.5.5. Cross validation and hyperparameter tuning

Nested cross-validation was used to develop and validate ML models and set the hyperparameter for ML algorithms. As depicted in Fig 1, the approach involves a two-level cross-validation structure. The outer cross-validation loop is responsible for estimating the generalization performance of the model. In each iteration of this outer loop, one data fold (out of ten) was held out as the test set, while the remaining folds were used for training and hyperparameter selection. Within the training data of each outer fold, an inner cross-validation loop was conducted to perform hyperparameter tuning. This involves partitioning the training data into five-folds, where different combinations of hyperparameters are assessed based on their performance across these inner validation folds. The hyperparameter configuration yielding the best average performance in the inner loop was selected for model training. Following the hyperparameter search, the model is retrained using the entire training data of the current outer fold with the identified hyperparameters. The trained model was then evaluated on the unseen test fold. This procedure is repeated across all outer folds (K=10, 10 times) to obtain an unbiased estimate of the model’s performance.

A grid search was employed for hyperparameter tuning to optimize model performance in the inner loop. This method systematically explores a predefined set of hyperparameter values by evaluating all possible combinations to identify the best configuration. The implementation was carried out using Scikit-learn’s GridSearchCV, automating the tuning process and ensuring efficient evaluation of multiple hyperparameter settings. In all inner loops, the optimal hyperparameters were found to be similar. Table S1 in supplementary material provide the details of parameters of each ML model.

#### 2.5.6. Evaluation metrics

Model performance was assessed using four standard classification metrics: accuracy, sensitivity, specificity, and area under the receiver operating characteristic curve (AUC). Accuracy represents the proportion of correctly predicted instances out of all predictions. Sensitivity (true positive rate) quantifies the model’s ability to correctly identify positive cases, while specificity (true negative rate) measures its capacity to accurately detect negative cases. AUC provides a threshold-independent summary of the model’s discriminative ability across all possible classification thresholds.

### 2.6. Explainability

To better interpret the importance of physical activity, sedentary behavior, and sleep features for detecting depression and anxiety symptoms, feature importance was assessed using SHapley Additive exPlanations (SHAP)^70^. SHAP is a widely used technique for interpreting the importance of features used in ML modeling after training. SHAP values quantify the impact of each feature on the output, providing a transparent ranking of their influence. SHAP values are calculated based on perturbations to the input data, estimating the marginal contribution of each feature by comparing predictions with and without it. This approach leverages game theory to fairly attribute predictive power across all features. The final values are indicative of feature importance, with higher values representing greater influence on the model’s output. We visualized the SHAP values for the 20 most influential features based on the best performing model.

## 3. RESULTS

### 3.1 Eligible participants and descriptive statistics

From a total of 5,861 cohort members who participated in the follow-up study and accepted to wear the accelerometer, valid accelerometer data were obtained from 4,006 individuals using the Hookie device and from 3,698 individuals using the Polar Active monitor. Among these, 2,810 participants provided valid data from both devices. After outlier detection, removal of faulty data, and merging with valid questionnaire responses, the final analytic samples included 1,825 participants for the BDI-II, 1,879 for the GAD-7, and 1,807 for the HSCL-25. Figure 1S in supplementary materials depicts the selection of eligible participants.

Full descriptive statistics of the study population and analytic groups are presented in Table 2. Compared to the total sample of participants with valid data from both accelerometers (N = 2810), the analytic group for the BDI-II (N = 1825) included a slightly higher proportion of females (66.0% vs 59.6%), employed individuals (86.9% vs 83.7%), and those with a university degree (29.6% vs 26.8%). Mean (SD) sleep duration in this subgroup was 451.10 (51.94) minutes per day, and average time spent on sedentary behavior, LPA, and MVPA were 439.24 (82.42), 348.51 (76.74), and 44.07 (21.45) minutes per day, respectively.

**Table 2:**
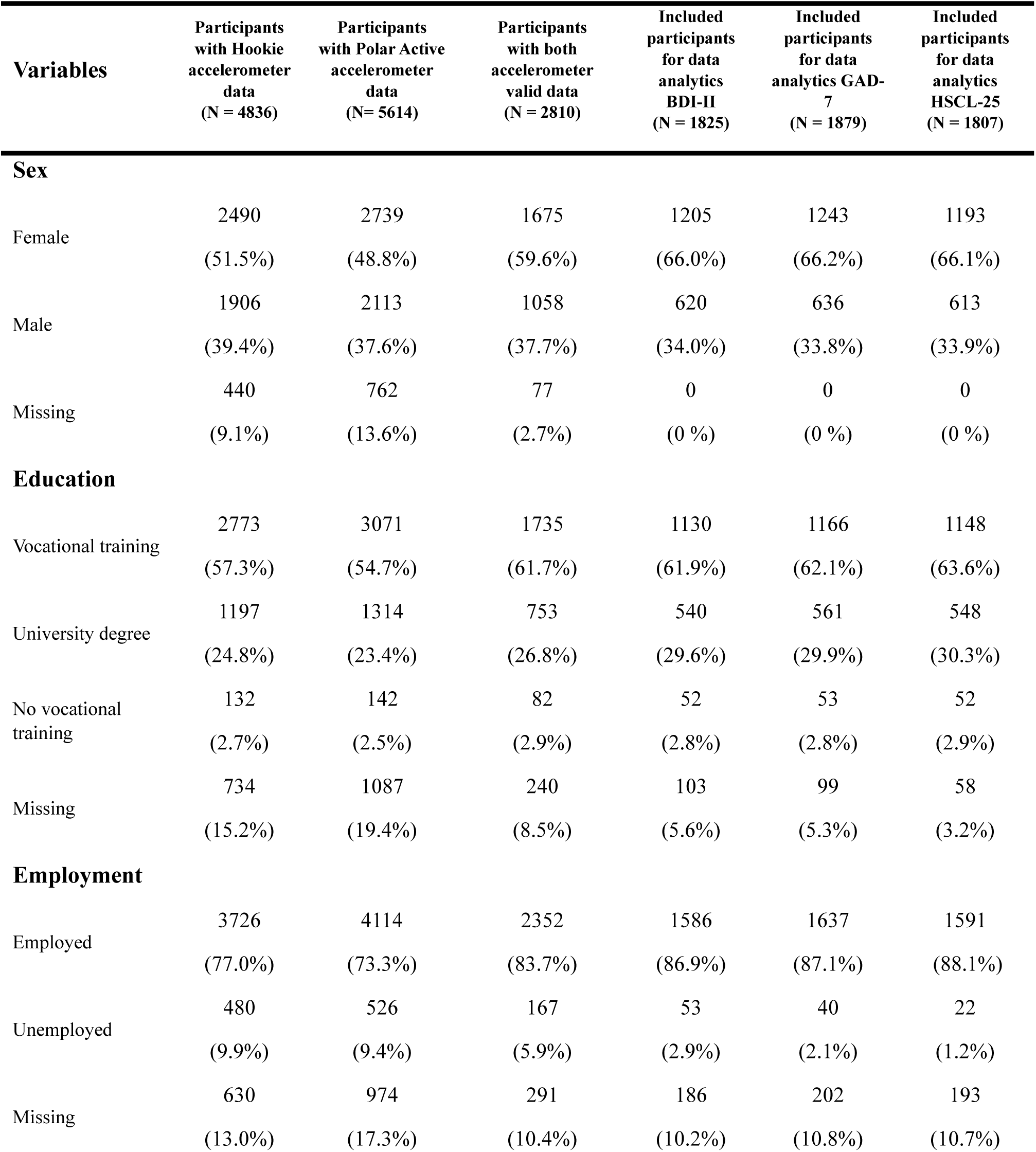
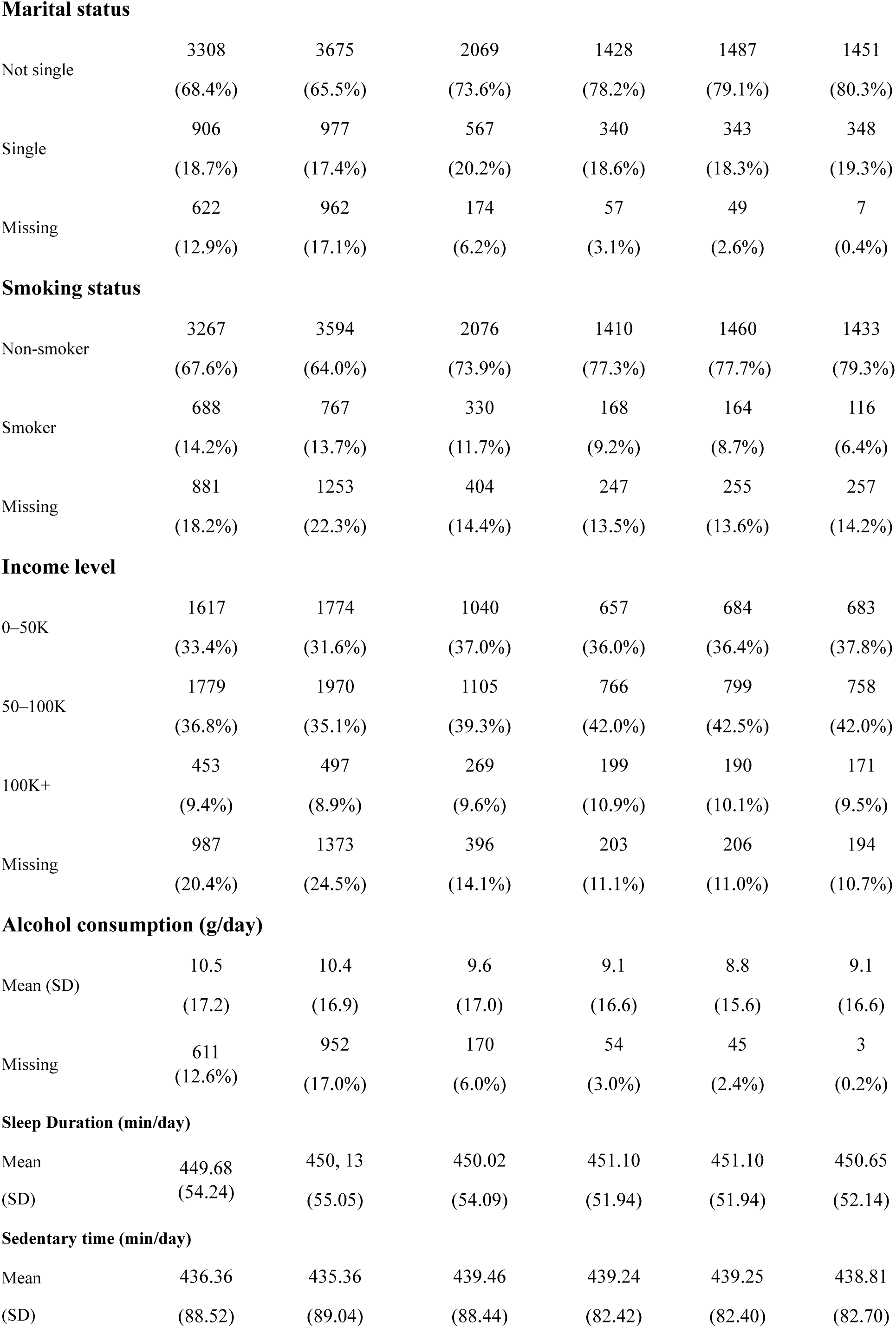
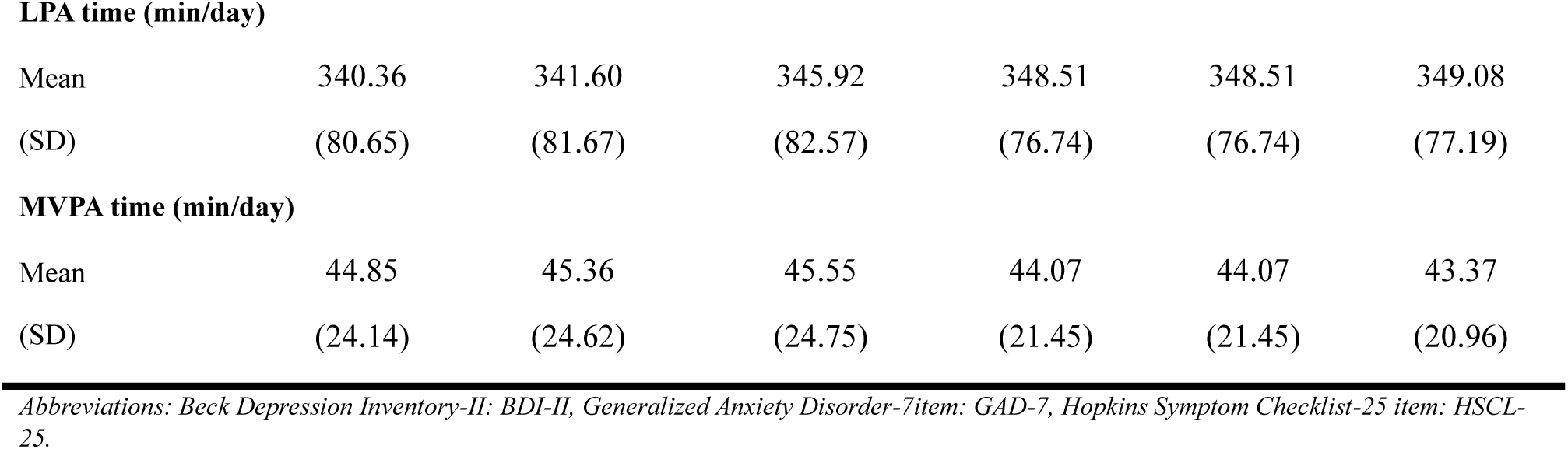
Characteristics and descriptive statistics of participants in this study.

| Variables | Participants with Hookie accelerometer data (N = 4836) | Participants with Polar Active accelerometer data (N = 5614) | Participants with both accelerometer valid data (N = 2810) | Included participants for data analytics BDI-II (N = 1825) | Included participants for data analytics GAD-7 (N = 1879) | Included participants for data analytics HSCL-25 (N = 1807) |
| --- | --- | --- | --- | --- | --- | --- |
| **Sex** |  |  |  |  |  |  |
| Female | 2490<br>(51.5%) | 2739<br>(48.8%) | 1675<br>(59.6%) | 1205<br>(66.0%) | 1243<br>(66.2%) | 1193<br>(66.1%) |
| Male | 1906<br>(39.4%) | 2113<br>(37.6%) | 1058<br>(37.7%) | 620<br>(34.0%) | 636<br>(33.8%) | 613<br>(33.9%) |
| Missing | 440<br>(9.1%) | 762<br>(13.6%) | 77<br>(2.7%) | 0<br>(0 %) | 0<br>(0 %) | 0<br>(0 %) |
| **Education** |  |  |  |  |  |  |
| Vocational training | 2773<br>(57.3%) | 3071<br>(54.7%) | 1735<br>(61.7%) | 1130<br>(61.9%) | 1166<br>(62.1%) | 1148<br>(63.6%) |
| University degree | 1197<br>(24.8%) | 1314<br>(23.4%) | 753<br>(26.8%) | 540<br>(29.6%) | 561<br>(29.9%) | 548<br>(30.3%) |
| No vocational training | 132<br>(2.7%) | 142<br>(2.5%) | 82<br>(2.9%) | 52<br>(2.8%) | 53<br>(2.8%) | 52<br>(2.9%) |
| Missing | 734<br>(15.2%) | 1087<br>(19.4%) | 240<br>(8.5%) | 103<br>(5.6%) | 99<br>(5.3%) | 58<br>(3.2%) |
| **Employment** |  |  |  |  |  |  |
| Employed | 3726<br>(77.0%) | 4114<br>(73.3%) | 2352<br>(83.7%) | 1586<br>(86.9%) | 1637<br>(87.1%) | 1591<br>(88.1%) |
| Unemployed | 480<br>(9.9%) | 526<br>(9.4%) | 167<br>(5.9%) | 53<br>(2.9%) | 40<br>(2.1%) | 22<br>(1.2%) |
| Missing | 630<br>(13.0%) | 974<br>(17.3%) | 291<br>(10.4%) | 186<br>(10.2%) | 202<br>(10.8%) | 193<br>(10.7%) |
| **Marital status** |  |  |  |  |  |  |
| Not single | 3308<br>(68.4%) | 3675<br>(65.5%) | 2069<br>(73.6%) | 1428<br>(78.2%) | 1487<br>(79.1%) | 1451<br>(80.3%) |
| Single | 906<br>(18.7%) | 977<br>(17.4%) | 567<br>(20.2%) | 340<br>(18.6%) | 343<br>(18.3%) | 348<br>(19.3%) |
| Missing | 622<br>(12.9%) | 962<br>(17.1%) | 174<br>(6.2%) | 57<br>(3.1%) | 49<br>(2.6%) | 7<br>(0.4%) |
| **Smoking status** |  |  |  |  |  |  |
| Non-smoker | 3267<br>(67.6%) | 3594<br>(64.0%) | 2076<br>(73.9%) | 1410<br>(77.3%) | 1460<br>(77.7%) | 1433<br>(79.3%) |
| Smoker | 688<br>(14.2%) | 767<br>(13.7%) | 330<br>(11.7%) | 168<br>(9.2%) | 164<br>(8.7%) | 116<br>(6.4%) |
| Missing | 881<br>(18.2%) | 1253<br>(22.3%) | 404<br>(14.4%) | 247<br>(13.5%) | 255<br>(13.6%) | 257<br>(14.2%) |
| **Income level** |  |  |  |  |  |  |
| 0–50K | 1617<br>(33.4%) | 1774<br>(31.6%) | 1040<br>(37.0%) | 657<br>(36.0%) | 684<br>(36.4%) | 683<br>(37.8%) |
| 50–100K | 1779<br>(36.8%) | 1970<br>(35.1%) | 1105<br>(39.3%) | 766<br>(42.0%) | 799<br>(42.5%) | 758<br>(42.0%) |
| 100K+ | 453<br>(9.4%) | 497<br>(8.9%) | 269<br>(9.6%) | 199<br>(10.9%) | 190<br>(10.1%) | 171<br>(9.5%) |
| Missing | 987<br>(20.4%) | 1373<br>(24.5%) | 396<br>(14.1%) | 203<br>(11.1%) | 206<br>(11.0%) | 194<br>(10.7%) |
| **Alcohol consumption (g/day)** |  |  |  |  |  |  |
| Mean (SD) | 10.5<br>(17.2) | 10.4<br>(16.9) | 9.6<br>(17.0) | 9.1<br>(16.6) | 8.8<br>(15.6) | 9.1<br>(16.6) |
| Missing | 611<br>(12.6%) | 952<br>(17.0%) | 170<br>(6.0%) | 54<br>(3.0%) | 45<br>(2.4%) | 3<br>(0.2%) |
| **Sleep Duration (min/day)** |  |  |  |  |  |  |
| Mean (SD) | 449.68<br>(54.24) | 450.13<br>(55.05) | 450.02<br>(54.09) | 451.10<br>(51.94) | 451.10<br>(51.94) | 450.65<br>(52.14) |
| **Sedentary time (min/day)** |  |  |  |  |  |  |
| Mean (SD) | 436.36<br>(88.52) | 435.36<br>(89.04) | 439.46<br>(88.44) | 439.24<br>(82.42) | 439.25<br>(82.40) | 438.81<br>(82.70) |
| **LPA time (min/day)** |  |  |  |  |  |  |
| Mean | 340.36 | 341.60 | 345.92 | 348.51 | 348.51 | 349.08 |
| (SD) | (80.65) | (81.67) | (82.57) | (76.74) | (76.74) | (77.19) |
| **MVPA time (min/day)** |  |  |  |  |  |  |
| Mean | 44.85 | 45.36 | 45.55 | 44.07 | 44.07 | 43.37 |
| (SD) | (24.14) | (24.62) | (24.75) | (21.45) | (21.45) | (20.96) |

Abbreviations: Beck Depression Inventory-II: BDI-II, Generalized Anxiety Disorder-7item: GAD-7, Hopkins Symptom Checklist-25 item: HSCL-25.

The analytic group for the GAD-7 (N = 1879) showed similar characteristics, with a slightly higher proportion of females (66.2% vs 59.6%), employed individuals (87.1% vs 83.7%), and participants holding a university degree (29.9% vs 26.8%) compared to the total sample. Mean (SD) sleep duration was 451.10 (51.94) minutes per day, with sedentary time, LPA, and MVPA averaging 439.25 (82.40), 348.51 (76.74), and 44.07 (21.45) minutes per day, respectively.

Similarly, the HSCL-25 analytic group (N = 1807) included a slightly higher proportion of females (66.1% vs 59.6%), employed participants (88.1% vs 83.7%), and those with a university degree (30.3% vs 26.8%). Participants mean (SD) sleep duration was 450.65 (52.14) minutes per day, with sedentary time averaging 438.81 (82.70) minutes per day, LPA 349.08 (77.19) minutes per day, and MVPA 43.37 (20.96) minutes per day.

### 3.2 Absence versus presence of depression symptom assessed by BDI-II (cutoff point >13 for depression)

The RF and stacked ensemble models both achieved the highest classification accuracy for depression symptom prediction, each reaching 70%. These two models also had the highest AUC scores, both reported at 70%. In terms of sensitivity, RF and stacked ensemble both reached 62%. The sensitivity of these models was 62%. The RF model had the highest specificity at 71%, meaning it was most effective at identifying individuals without depression symptoms. Other models including GBT, LightGBM, and XGB showed slightly lower performance, with accuracy scores ranging from 65% to 66% and AUC values between 66% and 68%. LR had the lowest accuracy at 56% and AUC 57% among all models tested (See Table 3).

**Table 3:** Performance of machine learning models predicting depression and/or anxiety symptom, as assessed by the BDI-II (cutoff point > 13), GAD-7 (cutoff point > 4), and HSCL-25.

| Risk score | Algorithms | Performance metrics |  |  |  |
| --- | --- | --- | --- | --- | --- |
|  |  | Accuracy,<br>% | AUC,<br>% | Sensitivity,<br>% | Specificity,<br>% |
| **Beck Depression Inventory-II (BDI-II)** | Random Forest (RF) | **70** | **70** | **62** | **71** |
|  | Extreme Gradient Boosting (XGB) | 66 | 68 | 60 | 65 |
|  | Logistic Regression (LR) | 56 | 57 | 51 | 58 |
|  | Gradient Boosting Tree (GBT) | 66 | 68 | 61 | 66 |
|  | Light Gradient Boosting Machine (Light-GBM) | 65 | 68 | **62** | 65 |
|  | Stacked Ensemble | **70** | **70** | **62** | 66 |
| **Generalized Anxiety Disorder-7item (GAD-7)** | Random Forest (RF) | **67** | **67** | 51 | **74** |
|  | Extreme Gradient Boosting (XGB) | 65 | 65 | 55 | 65 |
|  | Logistic Regression (LR) | 55 | 56 | 52 | 55 |
|  | Gradient Boosting Tree (GBT) | 64 | 66 | 53 | 68 |
|  | Light Gradient Boosting Machine (Light-GBM) | 64 | 65 | 54 | 66 |
|  | Stacked Ensemble | **67** | **67** | **55** | 70 |
| **Hopkins Symptom Checklist-25 item (HSCL-25)** | Random Forest (RF) | **72** | **70** | 54 | **75** |
|  | Extreme Gradient Boosting (XGB) | 65 | 67 | **60** | 66 |
|  | Logistic Regression (LR) | 57 | 58 | 54 | 58 |
|  | Gradient Boosting Tree (GBT) | 65 | 67 | 59 | 68 |
|  | Light Gradient Boosting Machine (Light-GBM) | 64 | 67 | 59 | 65 |
|  | Stacked Ensemble | 69 | **70** | 59 | 70 |

### 3.3 Absence versus presence of anxiety symptom assessed by GAD-7 (cutoff point > 4 for anxiety)

For anxiety symptom detection with the GAD-7, the RF and stacked ensemble models showed the highest accuracy, each at 67%. Both models also had an AUC of 67%. The highest sensitivity was observed in the stacked ensemble and XGB models, both scoring 55%. RF showed the best performance in specificity at 74%. LightGBM and GBT also performed reasonably well, with accurate values between 64% and 65%, and AUC scores ranging from 65% to 66%. LR had the lowest performance with an accuracy of 55% and AUC of 56%. (See Table 3).

### 3.4 Absence versus presence of depression and anxiety symptoms assessed by HSCL-25 (Cutoff point >1.75 for depression and anxiety)

In predicting depression and anxiety symptoms with HSCL-25, the RF model outperformed others, achieved the highest accuracy at 72% and an AUC of 70%. The stacked ensemble model followed with an accuracy of 69% and an AUC of 70%. Sensitivity was highest for the XGB model at 60%. The RF model had the highest specificity at 75%. Other models including GBT and LightGBM showed accuracy values between 64% and 65%, with similar AUCs. LR had the lowest performance across all evaluation metrics.

### 3.5 Absence versus presence of depression symptom assessed by BDI-II (cutoff point >19 for depression)

When depression was defined by higher cutoff point in BDI-II score, similar patterns of results were observed. Among the ML models evaluated for detecting depression symptoms, RF algorithm achieved the highest accuracy at 66% and the highest AUC at 66%. The model also showed sensitivity at 62% and the highest specificity at 66%. The GBT model also performed well, with an accuracy of 64% and an AUC of 66%. It had a sensitivity of 61% and a specificity of 64%. Light-GBM achieved the highest sensitivity at 66%, although its overall accuracy (62%) and specificity (62%) were slightly lower than RF and GBT. The stacked ensemble and XGB models showed performance with accuracies of 63% and 61%, respectively. LR showed the lowest overall performance with an accuracy of 58% and an AUC of 60%. However, its sensitivity (58%) was comparable to other models (See Table 4).

**Table 4:**
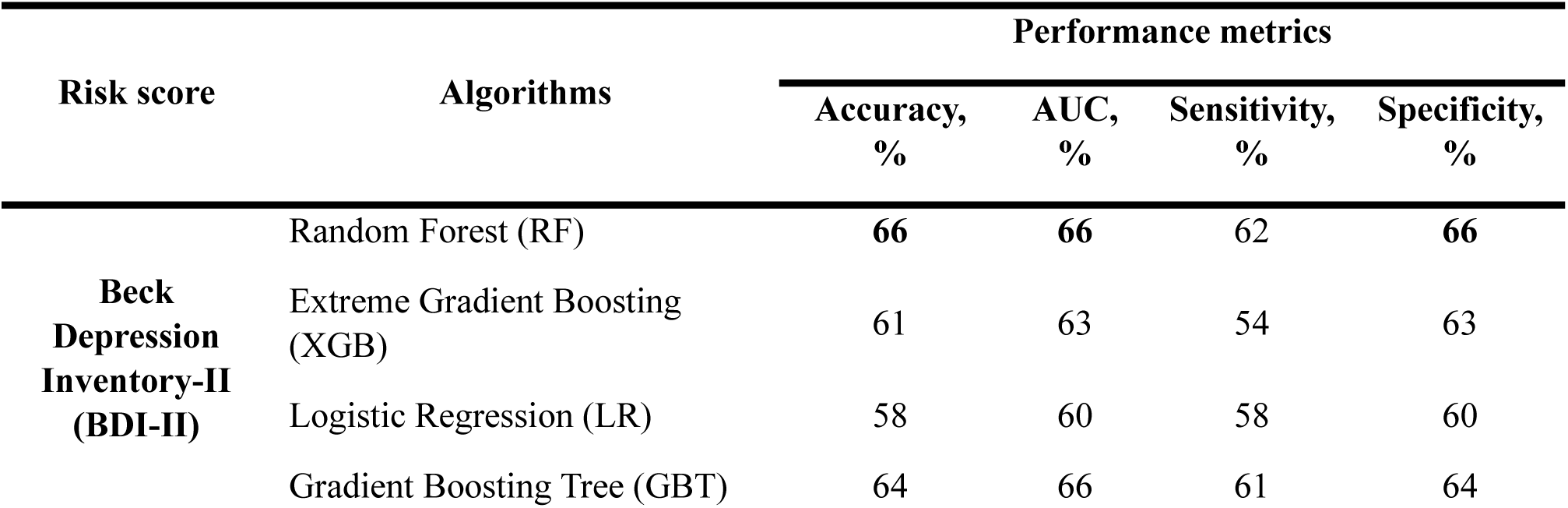
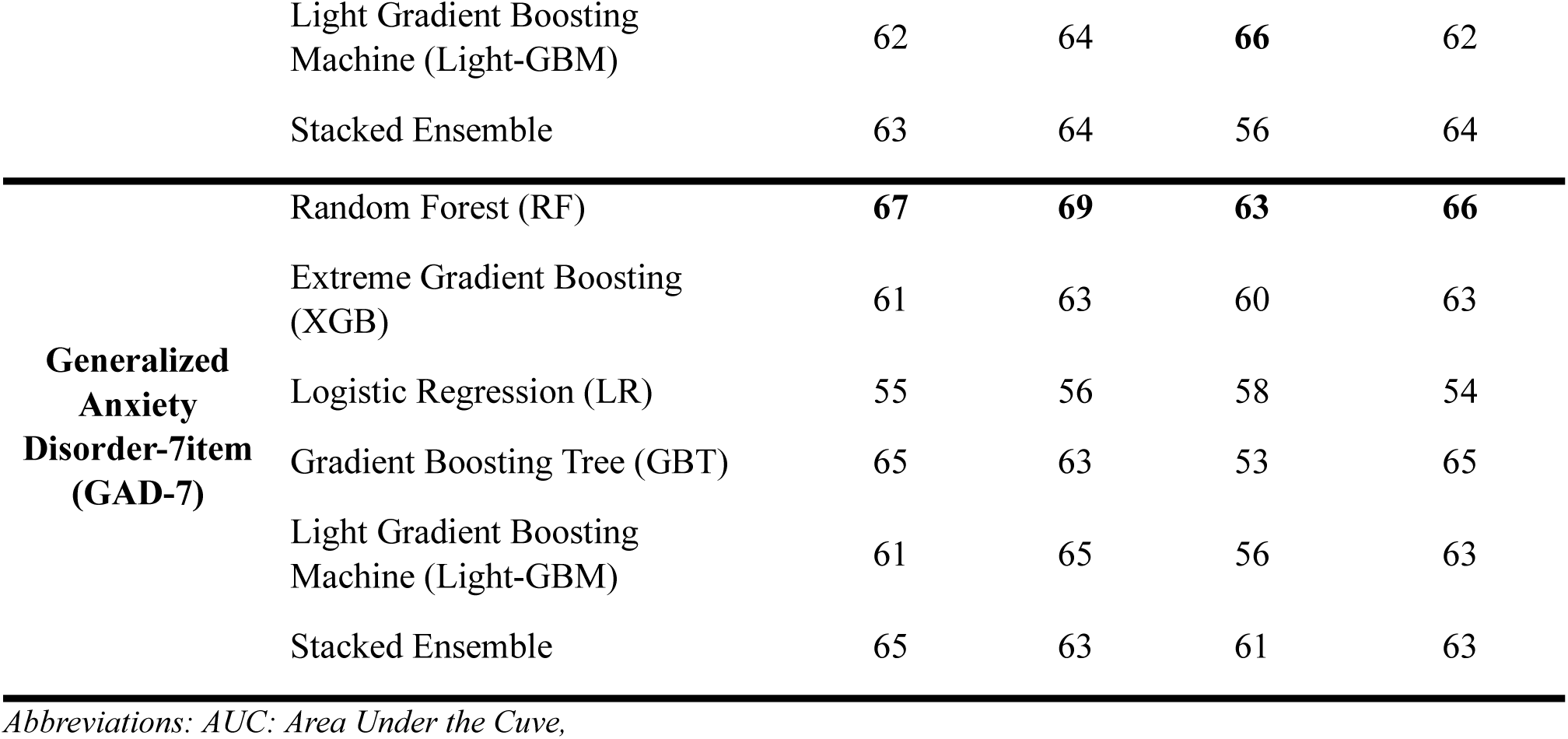
Performance of machine learning models applied to detect depression symptom and anxiety symptom based on BDI-II (cutoff point > 19) and GAD-7 (cutoff point > 9).

### 3.6 Absence versus presence of anxiety symptoms assessed by GAD-7 (cutoff point > 9 for anxiety)

When predicting anxiety symptom with considering the moderate and severe as anxiety group, the RF model achieved the highest performance across most metrics. RF reached an accuracy of 67% and the highest AUC at 69%. It also demonstrated a sensitivity of 63% and a specificity of 66%. The stacked ensemble and GBT models followed closely, each reporting an accuracy of 65%. The stacked ensemble model achieved a sensitivity of 61% and specificity of 63%, while GBT had a slightly lower sensitivity of 53% and a specificity of 65%. XGB and LightGBM models was performed with accuracy values of 61% and AUC scores between 63%–65%. Sensitivity values for these models ranged from 56% to 60%, and specificity values ranged from 63% to 65%. LR demonstrated the lowest performance across all metrics, with an accuracy of 55%, AUC of 56%, sensitivity of 58%, and specificity of 54% (See Table 4).

### 3.7 SHAP Explainability

Figure 2 illustrates the relative significance of physical activity, sedentary behaviors, and sleep-related features in predicting depression and anxiety symptoms, as assessed by BDI-II (cutoff point > 13), GAD-7 (cutoff point > 4), and HSCL-25 scores, according to SHAP values derived from the best-performing model (RF)

**Figure 2:**
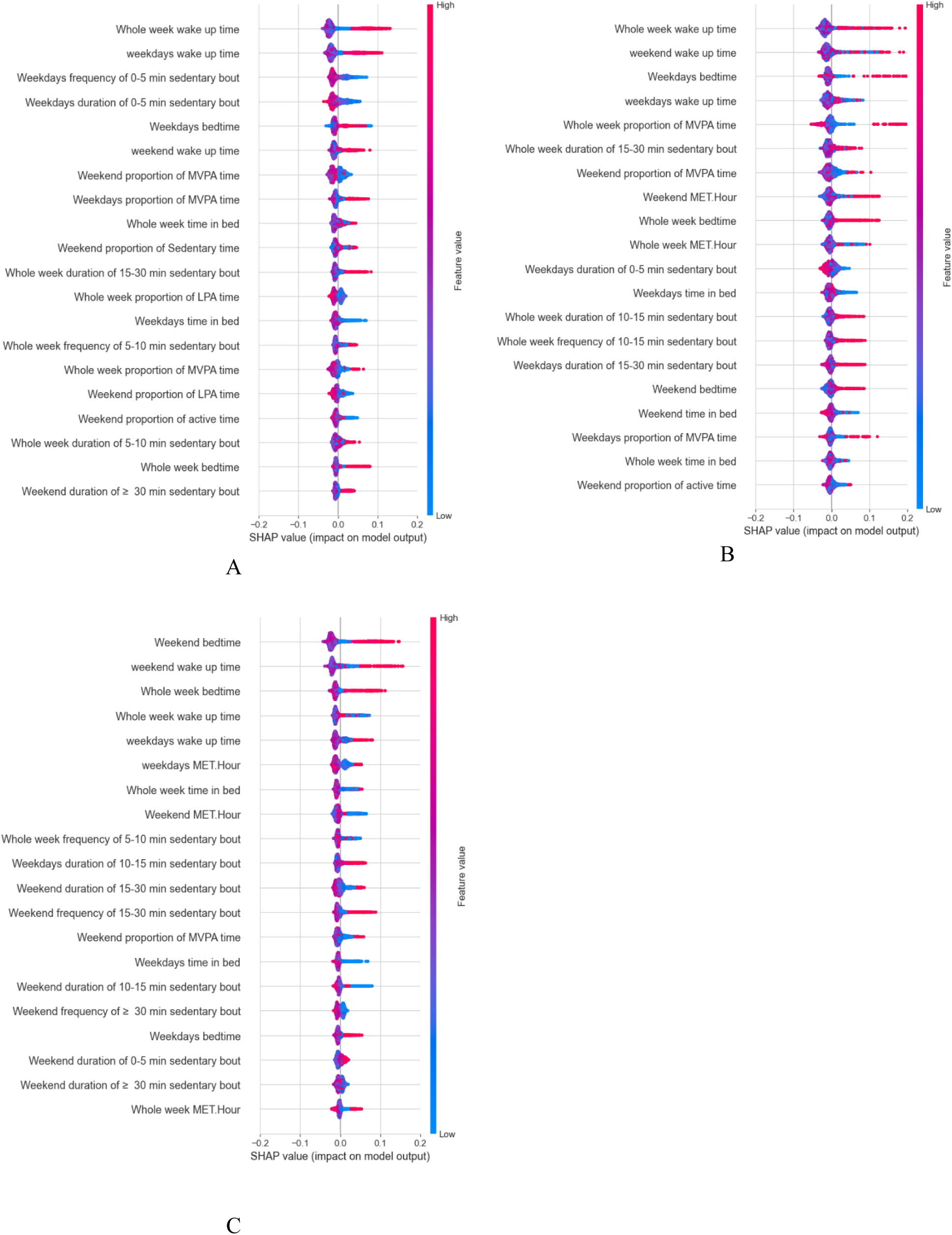
The plot shows how each feature affects the model’s prediction. The x-axis indicates the magnitude and direction of a feature’s influence on the model output. The color represents the feature value, with red indicating high values and blue indicating low values. Features are ranked in descending order of importance based on their average absolute SHAP value. A) Shows the interpretation of features in predicting depression symptom, B) Shows the interpretation of features in predicting Anxiety symptom, C) Shows the interpretation of features in predicting depression and anxiety symptom.

In Figure 2A, when depression symptoms were predicted using BDI-II scores, key predictive features for depression symptoms included *whole-week wake-up time*, *weekday wake-up time*, and the *frequency of 0–5-minute sedentary bouts*. Additional important variables included *weekday bedtime* and *weekend wake-up time*. Proportions of *MVPA* and *LPA* on both *weekdays* and *weekends* ranked highly in feature importance. SHAP values also indicated that a higher proportion of overall weekly sedentary behavior, as well as longer sedentary episodes lasting ≥30 minutes, significantly elevated predictions for depressive symptoms.

According to Figure 2B, when anxiety symptoms were predicted by GAD-7, *whole-week wake-up time*, *weekend wake-up time*, and *weekday bedtime* emerged as the most impactful features for detecting the presence of anxiety symptoms. *Whole-week MET hours* also contributed to the model for prediction of anxiety symptoms. *Weekend* and *weekday* proportions of *MVPA* appeared in the top 20 features. However, sedentary behavior variables, particularly the duration and frequency of sedentary bouts, had a mixed contribution.

In Figure 2C, when depression and anxiety symptoms were predicted using HSCL-25, the top predictive features included *weekend bedtime*, *wake-up times* (across both weekdays and weekends), and *whole-week bedtime*. Sedentary behavior features, particularly the *duration of 15–30-minute sedentary bouts*, also had relevant predictive value.

## 4. DISCUSSION

This population-based study developed and validated ML models to distinguish individuals with and without depression and anxiety symptoms using wearable accelerometer-determined physical behaviors metrics. Depression and anxiety symptoms were assessed with three widely used self-reported instruments. Among the ML algorithms evaluated, RF achieved the highest predictive ability to distinguish individuals with and without depression and anxiety symptoms. According to post analysis of interpretability of contributions of features, sleep-related features, particularly wake-up time, bedtime, and total time in bed, emerged as the most influential predictors of depression and anxiety symptoms. Wearable-determined physical activity and sedentary behaviors metrics showed to have only some information for detecting depression and anxiety symptoms. The findings suggest that depression and anxiety symptoms may be reflected in physical behaviors measured by wearable accelerometers, and that ML models can distinguish individuals with high symptom levels from those without.

In today’s digital age, a diverse array of data types such as blog posts, social media activity, and sensor signals and data collected from smartphones and wearable fitness trackers have been leveraged to identify individuals exhibiting symptoms of depression and anxiety. Systematic reviews have consistently reported that ML techniques applied to such data achieve classification accuracies ranging from 70% to 80% when distinguishing individuals with and without depression and anxiety^71,72^. In some cases, even higher accuracy is observed, depending on the data complexity and sophistication of the ML models used^72,73^. Our study could achieve a comparable level of accuracy reported in most studies using different digital data modalities to distinguish between individuals with and without depression and anxiety symptoms. Currently, it remains unclear which specific digital modalities offer the most predictive value for detecting depression and anxiety. Still, among all modalities, data generated from personal digital devices including smartphones and wearable activity and fitness trackers show a significant potential for digital phenotyping of mental health conditions. This potential stems from their widespread use, capacity for continuous data collection, and the ability to capture rich contextual information^14,74^.

Compared to smartphone-based digital phenotyping, which often relies on GPS traces, app usage, and communication patterns, wearable devices overall allow for passive and better measurement of behavioral and physiological signals. In particular, wearable accelerometry provides high-resolution insights without requiring active user engagement, thereby facilitating passive and unobtrusive data collection^10,75^. Wearable accelerometry is widely used for measurement of behavioral rhythms related to activity and rest cycles, physical activity and sedentary behaviors, and sleep behaviors^14,76^. While some studies have reported slightly better performance with smartphone-based digital phenotyping, primarily due to the richer contextual information captured, these methods face challenges such as inconsistent user interaction and privacy concerns^77^. In contrast, data derived from wearable devices tend to be more structured and less susceptible to such inconsistencies, although they may lack the broader behavioral and environmental context provided by smartphones^22,78^. In general, wearables offer an acceptable and scalable pathway for continuous monitoring and detection depression and anxiety symptoms, especially in general populations where unobtrusive data collection is critical^11,17,37^. Our findings support the utility of wearable-derived physical behavior digital phenotypes for the rapid detection of depression and anxiety symptoms.

Our findings suggest that depression and anxiety symptoms manifest in wearable accelerometer-determined physical behaviors in terms sleep timing features including wake-up time and bedtime, volume of physical activity, and higher sedentary time in relatively longer bouts. In particular, wake-up time, bedtime, and total time in bed, emerged as the most influential predictors of depression and anxiety symptoms. This aligns with growing evidence suggesting that disrupted rest-activity rhythms are closely associated with depression and anxiety symptoms and mood disorders. For instance, irregular sleep patterns and circadian misalignment have been linked to increased risk of depression and anxiety symptoms^79–81^. Recent studies further support this connection, showing that rest**-**activity rhythm metrics from wearable devices over 24-hour cycles are closely related to mental health^79,80^.

In our analysis, MVPA and MET-hours are relatively important predictors of depression and anxiety symptoms. This finding aligns with existing literature demonstrating an inverse relationship between physical activity levels and the risk of developing depression and anxiety disorders^82,83^. A systematic review found that higher levels of physical activity were associated with a 17% lower risk of depression and a 26% lower risk of anxiety^84^. While sedentary behaviors also contributed to prediction, it was generally less influential than activity and sleep related patterns. This supports prior evidence that prolonged sedentary periods are linked to higher rates of depression and anxiety^85^, but may have less predictive value when considered alongside activity and sleep related features.

Among the ML models evaluated, RF algorithm consistently outperformed others in distinguishing individuals with and without depression and anxiety symptoms. Despite its relatively strong performance, the overall moderate sensitivity observed across models highlights the need for further refinement. Future research should explore the integration of hybrid modeling strategies or deep learning approaches to improve sensitivity while maintaining acceptable specificity^10,36^. The findings of this study overall reinforce the potential utility of ML in combination with wearable accelerometer, potentially offering an objective, scalable, and non-invasive approach to screening for depression and anxiety symptoms^17^.

### 4.1 Strengths and limitations

Our study distinguishes itself not only by application of digital phenotyping to detect depression and anxiety symptoms from wearable-measured physical behaviors but also in its emphasis on methodological interpretability, an important aspect often overlooked in prior research on depression and anxiety symptoms prediction^10,17,36^. This study used data from a large population-based cohort study that included assessment of physical behaviors from wrist- and waist-worn accelerometers and mental health information. To date, several screening tools have been employed separately in previous studies examining depression or anxiety. We uniquely utilized three validated screening tools (BDI-II, GAD-7, and HSCL-25). Our ML analysis demonstrated comparable results independent of the specific screening tools employed (GAD-7, BDI-II, and the HSCL-25 Symptom Checklist). This study also utilized validated methods to measure physical activity intensities, sedentary behavior, and sleep patterns ^44^.

Our study is not without limitations. One of the limitations of this study is the use of two separate wearable devices to measure physical activity, sedentary behavior, and sleep. This may have increased participant burden and affected compliance. While we employed well-established self-report measures BDI-II, GAD-7, and HSCL-25 to assess symptoms of depression and anxiety, these tools function primarily as screening instruments rather than diagnostic tools. Although widely used in epidemiological and population-based research^48,50^, these measures do not offer the diagnostic precision of structured clinical interviews, which are considered the gold standard in psychiatric assessment. Future research should explore whether the similar relationships observed between accelerometer-derived features and mental health outcomes when outcomes are defined through clinical diagnostic interviews.

## 5. CONCLUSION

This study highlights the promising potential of utilizing wearable accelerometer-determined physical behaviors with ML approaches to detect individuals with and without depression and anxiety symptoms in a general population. ML models trained on wearable accelerometer-determined physical behaviors metrics achieved reasonably good accuracy detecting individuals with and without depression and anxiety symptoms. Analysis of contribution features revealed that while wearable accelerometer-determined sleep-related metrics are among the most influential features, other physical behavior metrics including physical activity and sedentary behavior metrics also have some information for predicting depression and anxiety symptoms. This suggests that depression and anxiety may manifest in wearable accelerometer-determined physical behaviors and using ML models with wearable accelerometer-determined physical behaviors metrics could potentially be a promising method for scalable and rapid screening of depression and anxiety in a general population.

## Data Availability

NFBC data is available to qualified researchers by applying through the University of Oulu's NFBC project center.

## Acknowledgements

VF is supported by TU Dortmund university. The present study is connected to the DigiHealth and 6GESS strategic profiling projects at the University of Oulu supported by the Research Council of Finland (project number 326291, 336449) and the University of Oulu. This study has also received funding from the Ministry of Education and Culture in Finland [grant numbers OKM/20/626/2022, OKM/76/626/2022, OKM/68/626/2023]. The funders played no role in designing the study; collecting, analyzing, and interpreting the data; or writing the manuscript. The results of the study are presented honestly and without fabrication, falsification, or inappropriate data manipulation.

## Declarations

The authors declare that there is no conflict of interest.

## Supplementary materials

**Table S5.** Hyperparameter tuning of algorithms.

| Algorithm | Best Parameters |
| --- | --- |
| **Random forest (RF)** | 'n_estimators': [300],<br>'max_depth': [10],<br>'min_samples_split': [5],<br>'random_state': [42] |
| **Extreme gradient boosting (XGBoost)** | 'n_estimators': [300],<br>'learning_rate': [0.05],<br>'max_depth': [5],<br>'eval_metric': ['logloss'], |
| **Logistic regression (LR)** | 'max-iter': [1000],<br>'kernel': [rbf], |
| **Gradient Boosting Tree (GBT)** | 'n_estimators': [300],<br>'learning_rate': [0.1],<br>'max_depth': [5],<br>'random_state': [42] |
| **Light gradient boosting machine (LightGBM)** | 'n_estimators': [300],<br>'learning_rate': [0.05],<br>'max_depth': [10] |

**Figure S1:**
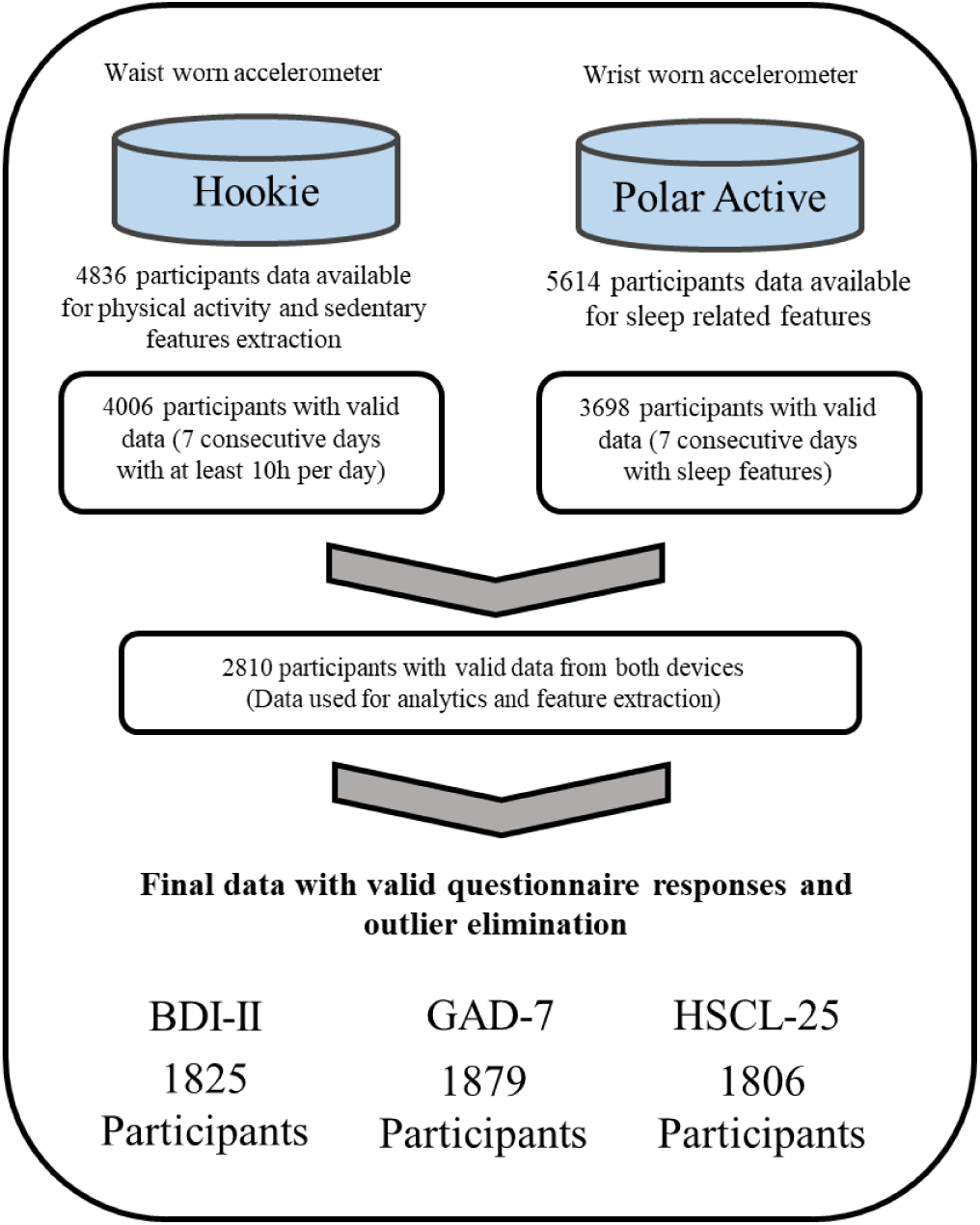
Flow of included participants (NFBC1966) data for this study.

